# Is climate a primary driver of Vietnam’s dengue, or a shared long-term trend? A five-method analysis over 36 years

**DOI:** 10.64898/2026.08.12.26360281

**Authors:** Doanh Nguyen-Ngoc, James M. Trauer, Hai Son Vo, Andrew W. Taylor-Robinson, Thanh H. Nguyen, Thi Ngan Vuong, Viet Long Bui

## Abstract

**Background:** Vietnam’s reported dengue burden has risen roughly five-fold since 1990. Multi-decadal studies linking climate indices to dengue rarely separate genuine year-to-year coupling from a long-term trend the two share.

**Methods:** We assembled a provenance-preserving national annual dengue series (1990-2025; OpenDengue plus Ministry of Health figures) and correlated it with annual and March-May means of eight tropical sea surface temperature (SST) indices at lags of zero and one year under five trend-correction lenses: raw, linear detrending, first-differencing, socio-demographic-index residualisation and AR(1) prewhitening.

**Findings:** Cases rose 3·5 percent annually. Seven predictors were significant at zero lag, led by the annual Indian Ocean Basin-Wide index (IOBW; Spearman +0·534), but linear detrending removed all. First-differencing preserved six, led by spring IOBW (+0·468), the annual Atlantic Multidecadal Oscillation (AMO; +0·462) and annual IOBW (+0·423); three survived AR(1) prewhitening - annual AMO and annual and spring IOBW. Spring AMO and a lag-1 Tropical North Atlantic signal (−0·452) did not, and are hypothesis-generating. El Niño-Southern Oscillation indices failed throughout.

**Interpretation:** Most of the apparent association reflects a trend shared by warming oceans and expanding surveillance; we could not demonstrate that climate is the primary driver at this scale. Trend is not the whole story: IOBW and annual AMO persist under trend- and persistence-removing transformations. Because transmission responds to climate over weeks to months, annual averaging smooths the lags through which El Niño acts; these nulls reflect temporal scale, not climate insensitivity; usable predictors will require monthly, province-level models.

**Funding:** Center for Environmental Intelligence, VinUniversity (project VUNI.CEI.FS_0001).

**Research in context:** *Evidence before this study:* We searched PubMed, Web of Science and Google Scholar for studies published up to May 2026 linking large-scale climate indices or sea surface temperature to dengue incidence, combining dengue, climate, sea surface temperature, ENSO, teleconnection and time-series terms with Vietnam, without language restriction. Many studies covering two or more decades reported strong correlations between basin-scale indices and national dengue counts. Most, however, relied on raw correlations or a single detrending choice, and rarely tested whether an apparent association reflected genuine year-to-year coupling or merely a shared long-term trend.

*Added value of this study:* Most long-term studies remove the shared upward trend in only one way, or not at all. To our knowledge this is the first study to compare five trend-correction methods on a multi-decadal national dengue record and to read their agreement or disagreement as a diagnostic of which climate signals are real. A signal that appears only before the trend is removed is probably following it; one that persists is more likely real. Applied to a record spanning more than three decades, this comparison separates the two: several widely reported raw correlations weakened once the shared trend was accounted for.

*Implications of all the available evidence:* Climate-informed analyses of multi-decadal data should report at least two trend-correction approaches alongside the raw correlation and treat their disagreement as evidence about where a signal sits, rather than operationalising raw long-span correlations. For Vietnam, the apparent national-scale association is dominated by a shared long-term trend but retains a smaller, robust inter-annual component led by the Indian Ocean and AMO signals; genuine coupling is more likely detectable at monthly resolution and provincial scale, where statistical power and physical mechanism are jointly available. Surveillance systems should retain explicit source provenance, so trend-corrected re-analysis remains possible as records grow.

## Introduction

Dengue is the most widespread arboviral disease globally, with approximately 390 million annual infections and 96 million symptomatic cases.^1^ Vietnam ranks among the most severely affected countries in the Western Pacific Region, with reported cases rising roughly five-fold over the 36-year record ^2^. These annual totals are highly variable, swinging several-fold between successive years. Identifying the drivers of this rising burden, including the extent to which global tropical sea surface temperature (SST) modes influence inter-annual variation, is a prerequisite for operational forecasting and climate-adaptive public health planning.

A substantial literature has examined climate-dengue relationships at global^3,4^, regional, and sub-national scales [6-8]. At the global scale, Chen and colleagues^3^ identified the Indian Ocean Basin-Wide (IOBW) index as the strongest single predictor of annual dengue incidence across 46 countries, outperforming El Niño-Southern Oscillation (ENSO) diagnostics. At the regional scale, Tian and colleagues attributed 63 percent of dengue variance in 57 countries to ENSO^8^. In Vietnam, Gibb and colleagues^5^ used hierarchical Bayesian models with provincial local climate covariates, while Colón-González and colleagues^6^ combined climate and Ministry of Health (MoH) surveillance data in a superensemble forecast. These studies have collectively advanced our understanding of climate drivers, but two methodological gaps persist.

First, a correlation computed directly from (raw or log-transformed) case counts and climate indices does not distinguish a shared long-term trend from year-to-year anomalies, and the two differ in what they imply for practice. Over three decades both tropical SSTs and Vietnam’s dengue surveillance counts have risen substantially - IOBW by about 0·5 degrees C from the 1990s to the 2020s, and reported annual cases by about 3·5 percent per year - such that any raw correlation may plausibly reflect this shared trend rather than genuine year-to-year coupling. A trend shared by climate, urbanisation, vector range expansion and improved case ascertainment cannot be attributed to climate from the correlation alone, and treating it as a climate signal may lead to misdirection of control resources.^5^ Whether such a rise constitutes real burden growth or is mainly attributable to population growth and wider improvements in surveillance coverage cannot be determined from case counts alone. Only the inter-annual component of a climate predictor - its year-to-year departures from the long-term trend, acting on Aedes vector density, the virus’s extrinsic incubation period, or population immunity - carries the information that could give genuine year-to-year predictive value; a predictor that tracks dengue only through a shared trend can look skillful in hindcast yet add little at the inter-annual scale. Annual resolution imposes a further limit of its own: because Aedes life cycles and viral incubation operate over weeks to months, calendar-year averaging smooths the three-to-six-month lags through which climate acts, so a null association at annual resolution constrains the scale at which a signal can be detected rather than establishing that dengue is insensitive to climate.

Second, a dengue series spanning several decades is rarely drawn from a single source. OpenDengue version 1.3assembles Vietnam’s 1990-2022 records from multiple underlying sources with differing coverage, temporal resolution and case definitions^2^. If a merged series does not record which dataset supplied each year, a shift caused by a source handover cannot be distinguished from a genuine change in dengue risk, and can pass unnoticed into attribution and forecasting conclusions. Records for 2023-2025, from public MoH communications, likewise need comparability checks against the earlier years.

Vietnam operates an Electronic Communicable Disease Surveillance (eCDS) system^9^, but it does not cover the pre-digital era and its historical line-list data are not publicly available, such that it cannot supply the full 1990-2025 record. As such, improving the reach and forecasting value of dengue surveillance still depends on a complete multi-decade series, which we assemble from openly available sources - OpenDengue version 1.3 for 1990-2022 and MoH communications for 2023-2025 - an approach that is also transparent and reproducible.

The present study addresses both gaps. We build a national annual dengue series for 1990-2025 from OpenDengue and MoH sources, applying a deterministic priority rule that preserves source provenance for each year. We report GBD modelled incidence and DALYs only as descriptive context, and test 16 climate predictors against 36 annual case totals with multiple-testing control across seven sensitivity configurations (including two trend-decomposition methods), to identify which climate-dengue associations are robust enough to be interpreted as genuine inter-annual coupling and which are artefacts of shared trends. Applying these five methods as a single panel, we show that raw multi-decadal correlations are dominated by shared trend, and we argue that reporting such a panel should become standard practice for climate-disease studies spanning more than two decades.

## Methods

### Study design and setting

We performed a descriptive and correlational analysis of Vietnam’s national reported dengue burden at annual resolution, 1990 to 2025 (36 years). Vietnam’s population grew from approximately 66 to 101 million over the analysis period. Vietnam’s dengue surveillance was progressively strengthened over this period, moving from paper-based provincial reporting toward electronic case reporting, although the exact dates of these changes are not consistently documented.

For 1990 to 2022, Vietnam’s dengue case data were drawn from the OpenDengue version 1.3 database^2^. For each year we extracted the national (country-total) record at the finest temporal resolution available and summed it to an annual total. Where a year was covered by more than one source, we selected a single source using a fixed order of preference: WHO Western Pacific Regional Office surveillance records first, then the Project Tycho database^11^, then peer-reviewed literature compilations, and finally Vietnamese MoH reports. This hierarchical approach identified a single unique source for every year. The full priority rule and the source assigned to each year are given in Supplementary Methods S1.1 and Supplementary Table S1, and the full data-extraction protocol in Supplementary Methods S1.2.

### Climate variables selection

We tested eight tropical SST indices spanning the Indian, Pacific, and Atlantic basins - chosen as the principal tropical sea-surface-temperature modes implicated in Southeast Asian monsoon variability and arboviral transmission, with El Niño-Southern Oscillation diagnostics included for comparability with earlier dengue studies (full rationale in Supplementary Methods S1): the Indian Ocean Basin-Wide index (IOBW) and Dipole Mode Index (DMI); the Multivariate ENSO Index (MEI), Oceanic Niño Index (ONI), Southern Oscillation Index (SOI), and Pacific Decadal Oscillation (PDO); and the Atlantic Multidecadal Oscillation (AMO) and Tropical North Atlantic index (TNA). Each was aggregated to an annual mean and a March-May spring mean and tested at lags of zero and one year, giving 16 candidate predictors against 36 annual dengue observations (1990-2025). Definitions, data sources, computation, and selection rationale are given in Supplementary Methods S1, and the annual and spring series of all eight indices are plotted in Supplementary Figure S1.

### Statistical analysis

Long-term trend was assessed using the non-parametric Mann-Kendall test^12,13^ with tie-corrected variance and Sen’s slope estimator. A parametric log-linear regression of the natural logarithm of annual case counts on calendar year provided an annual percentage-change estimate. Outbreak and trough years were identified from the detrended log-transformed series as years lying more than 1·5 standard deviations above the mean (outbreak years) or below it (trough years). Autocorrelation of the detrended log-transformed series was computed for lags of 1 to 12 years, with approximate 95 percent significance bounds for white noise set at plus or minus 1·96 divided by the square root of the number of years. Change-points were detected with the Pruned Exact Linear Time algorithm (PELT)^14^ using a radial basis cost function.

Associations between climate predictors and log-transformed annual dengue counts were quantified using Spearman rank correlation at two lags: no lag (same-year climate with same-year dengue) and a one-year lag (previous-year climate with the current year’s dengue). Benjamini-Hochberg false-discovery-rate control at the 5 percent level^15^ was applied separately within each lag across the 16 predictors, giving Benjamini-Hochberg-adjusted p-values. For each predictor we reported the stronger of the two lags (the larger correlation in absolute value) and ranked predictors accordingly (see Supplementary Methods S1.3 for further detail). Because the detrended case series shows no autocorrelation beyond lag 0 (Figure 2B), the zero-lag (same-year) analysis is treated as the primary analysis and one-year-lag results are reported as exploratory.

We applied five trend-correction transformations to the dengue and climate series before computing associations, each designed to test a distinct hypothesis about the source of any observed association: whether it reflects shared long-term trend, year-to-year interannual coupling, or confounding by socioeconomic development. The five transformations are described below; their agreement or disagreement across analyses is itself informative: rather than treating survival under every transformation as a pass/fail criterion, we interpret each association by the pattern of transformations it survives (for example, surviving first-differencing and AR(1) prewhitening but not linear detrending locates a signal in genuine inter-annual variation that remains partly entangled with the shared trend).

Specifically: (i) raw correlation tests whether dengue and climate co-vary across the full 36-year window, including any shared multidecadal trend; (ii) linear detrending tests whether deviations from each series’ fitted linear trend are correlated; (iii) SDI residualisation removes the development-related component shared by both series, testing whether the association persists after controlling for socioeconomic confounding; (iv) AR(1) prewhitening removes short-term serial autocorrelation from each series; (v) first-differencing replaces each series with its year-on-year change, removing all shared long-term trend and isolating interannual coupling.

### Primary and sensitivity analyses

First-differencing of the log-transformed case series and each climate predictor was treated as the primary trend-correction approach because it removes shared long-term trend and isolates inter-annual coupling, the construct of interest at the national annual scale. The four other approaches (raw correlation, linear detrending, SDI residualisation and AR(1) prewhitening) are reported as sensitivity analyses. A predictor whose association did not retain its sign and rough magnitude under first-differencing was not interpreted as evidence of inter-annual climate-dengue coupling, only of shared long-term co-trend. The 16-predictor ranking was repeated under seven sensitivity configurations. Four varied the years included: the full 36-year series, and series excluding 2011 (a single-source year), the COVID era of 2020 to 2022, and the MoH-only years of 2023 to 2025 - leaving 35, 33, and 31 years respectively. Three applied trend correction before re-correlating: linear detrending of both the log-transformed cases and each predictor; first-differencing of the two series; and residualisation of the log-transformed cases on Vietnam’s SDI (quadratic fit). The last is a domain-informed alternative to purely statistical detrending - a predictor that survives if it cannot be attributed to a socio-economic development trend. The two trend-correction transforms are defined in Supplementary Methods S1.4, and the Spearman correlation under every configuration is tabulated in Supplementary Table S3.

Separately, as the fifth transformation, the log-transformed case series was AR(1)-prewhitened to remove first-order serial dependence before re-correlation (Supplementary Methods S1.5), with the resulting correlations for the surviving predictors presented in Supplementary Table S4.

### IOBW-focused inference

For the primary IOBW finding, we regressed the log-transformed case counts on spring IOBW at zero lag by ordinary least squares and took the slope, with a 95 percent confidence interval from 5,000 bootstrap resamples. The slope and its bootstrap confidence interval were also computed for the first-differenced version of the two series. The two scatter relationships are shown in Supplementary Figure S3, and the bootstrap slope across 13 specifications in Supplementary Figure S4. AR(1) prewhitening with a percentile bootstrap confidence interval was applied to all six predictor-lag combinations that survived first-differencing, rather than to a pre-selected subset; the full results are reported in Supplementary Table S4.

### Software and reproducibility

All analyses were performed in Python^16^. The analysis code that reproduces all results and figures will be released in a public GitHub repository: https://github.com/vlbui/dengue-climate-vietnam.

## Results

Reported dengue notifications rose from 37,569 cases in 1990 to 190,040 in 2025, with peaks in 1998 (234,866) and 2022 (367,729). The full series is shown in Figure 1. Full provenance is reported in Supplementary Table S1. For contextual reference, GBD 2023 modelled estimates indicate that Vietnam’s dengue burden is concentrated in children and young adults, with a gradual shift towards older age groups over the study period (Supplementary Figure S5).

**Figure 1.**
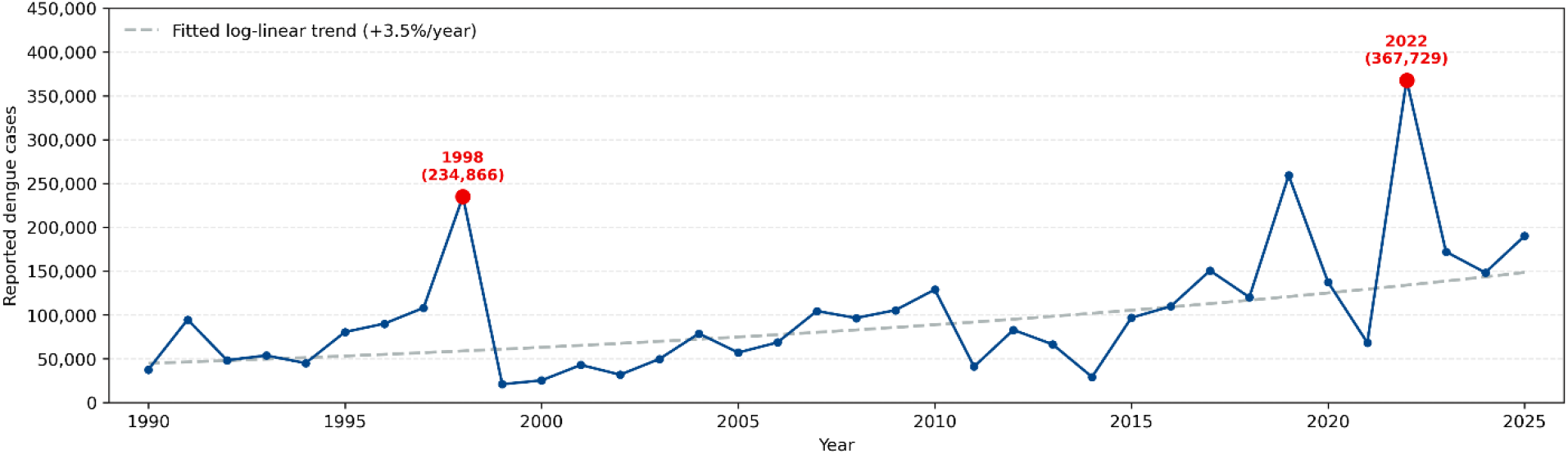
Vietnam’s annual reported dengue cases, 1990 to 2025, from OpenDengue version 1.3 merged with Ministry of Health figures for 2023 to 2025.

This increase was statistically significant (Mann-Kendall p < 0·001) but irregular: 1998 and 2022 stood out as outbreak years and 1999, 2000 and 2014 as trough years. The 2020-2021 COVID-19 disruption was associated with a notable dip in reported cases - 137,470 in 2020 and 68,268 in 2021, down from 259,070 in 2019 - before rebounding to the 2022 record high; even the 2021 low did not reach the trough-year threshold. The series showed no obvious regular multi-year cycle of the kind serotype rotation would produce (Figure 2), and its change-points did not coincide with the 2009, 2016 or 2020 milestones.

**Figure 2.**
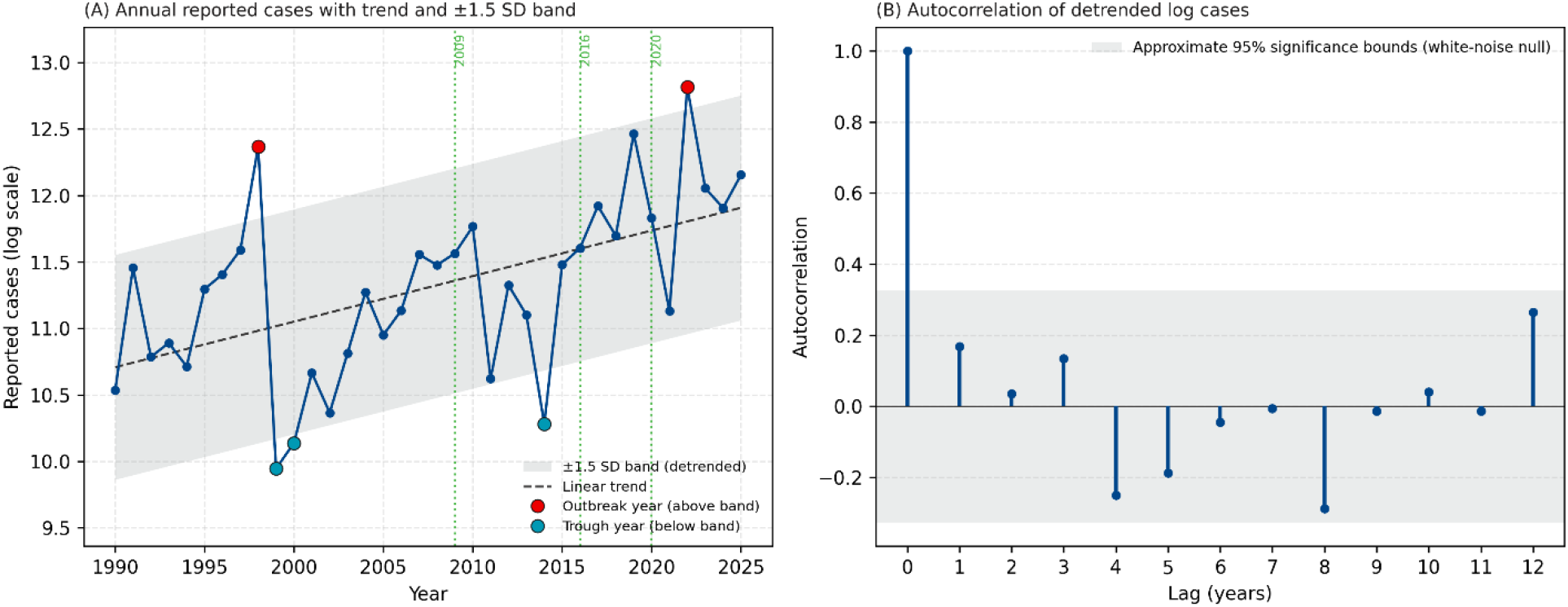
Two-panel annual descriptive summary of reported dengue in Vietnam, 1990 to 2025. (A) Annual reported cases on a log scale, shown with the fitted linear trend and a shaded band at plus or minus 1·5 standard deviations of the detrended series; the two outbreak years (1998 and 2022, red) lie above the band and the three trough years (1999, 2000 and 2014, teal) lie below it. Three dotted green vertical lines mark health-system milestones (the 2009 WHO dengue case-classification revision, the 2016 national electronic surveillance launch, and the 2020 COVID-19 disruption). (B) Autocorrelation of detrended log cases; the shaded band shows the approximate 95 percent significance bounds for white noise (no autocorrelation).

In raw correlations, seven of the sixteen climate predictors were significant at zero lag after multiple-testing correction (annual DMI was additionally significant at a one-year lag); all were Atlantic or Indian Ocean SST, led by the annual IOBW index (rank correlation +0·534). ENSO indices were weak and never significant (Figure 3); the full set of 16 correlation coefficients is given in Supplementary Table S2. Plotting the case record against all eight indices on a common standardised axis shows the same contrast visually: the Indian Ocean and Atlantic indices rise and fall with reported cases across the record, whereas the Pacific and ENSO indices do not (Figure 5).

**Figure 3.**
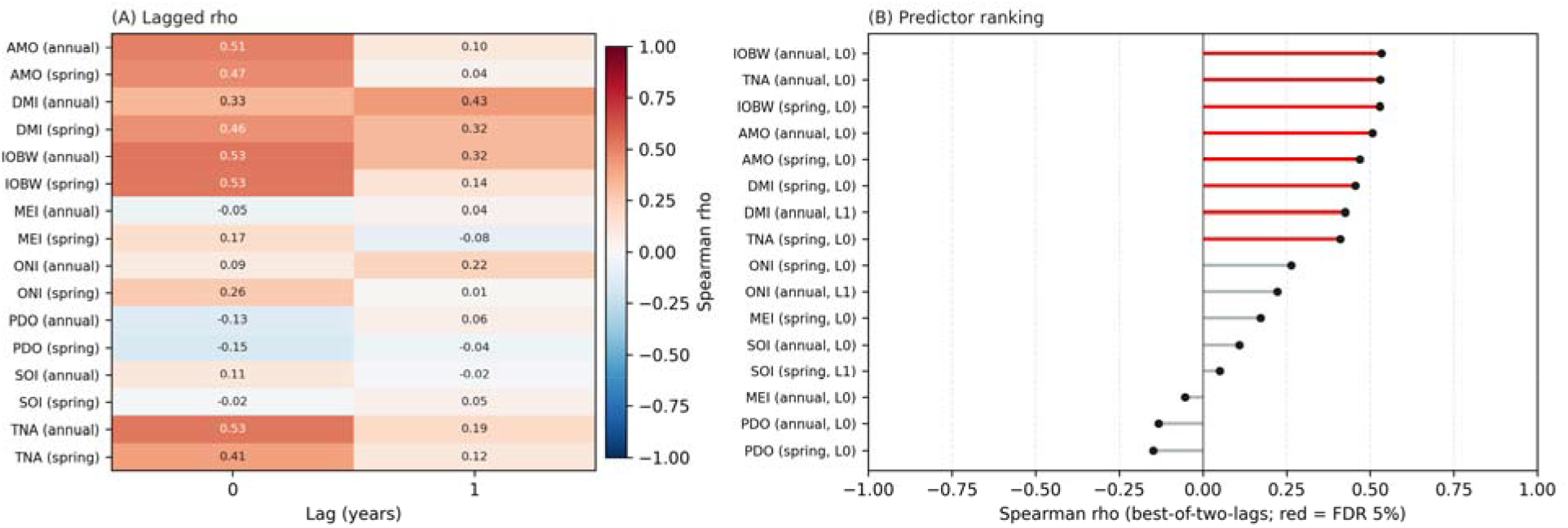
Raw climate-dengue correlations. (A) Heatmap of Spearman rho between ln(reported dengue cases) and 16 climate predictors (AMO, DMI, IOBW, MEI, ONI, PDO, SOI, TNA × annual and spring MAM means) at lag 0 and lag 1 years. (B) Lollipop ranking of each predictor’s best-of-two-lags rho; red entries are Benjamini-Hochberg FDR-significant at 5 percent.

These raw correlations, however, were not stable. When split by data source into the Project Tycho, WHO Western Pacific Regional Office and Ministry of Health periods, the leading predictors weakened considerably and none remained significant within any single source (Supplementary Figure S2). Spring IOBW illustrates this most clearly: its zero-lag rank correlation fell from +0·39 in the Project Tycho period to −0.02 (WHO WPRO) and +0·09 (Ministry of Health), while the annual IOBW index was not reproduced within any single source (+0·39, +0·05 and −0·31, respectively). Strong full-sample correlations combined with near-zero within-period correlations is the classic signature of an association driven by shared long-term trend rather than year-to-year coupling.

Under the primary transformation, first-differencing (Methods), signals in both the Indian and Atlantic basins survived; the leading Indian Ocean basin-wide signal retained its positive sign and significance, and under AR(1) prewhitening persisted alongside the annual AMO, and was therefore interpreted as evidence of inter-annual coupling. Trend correction confirmed this (Figure 4); the coefficient under each transformation is listed in Supplementary Table S3, with the AR(1)-prewhitened values in Supplementary Table S4. Linear detrending removed all seven signals, and the stricter SDI residualisation removed every signal. First-differencing preserved six signals spanning both ocean basins: annual and spring IOBW and annual and spring AMO at zero lag (all positive), and spring and annual TNA at a one-year lag (negative). Applying AR(1) prewhitening uniformly to all six (Supplementary Table S4) left three with a bootstrap 95% confidence interval excluding zero - annual AMO and both annual and spring IOBW - whereas the spring AMO and both TNA signals did not survive prewhitening. The leading raw signal, IOBW, therefore retained a positive inter-annual association under both the trend-removing (first-differencing) and persistence-removing (AR(1)) lenses, indicating a genuine year-to-year component rather than a coupling residing solely in the shared multidecadal trend.

**Figure 4.**
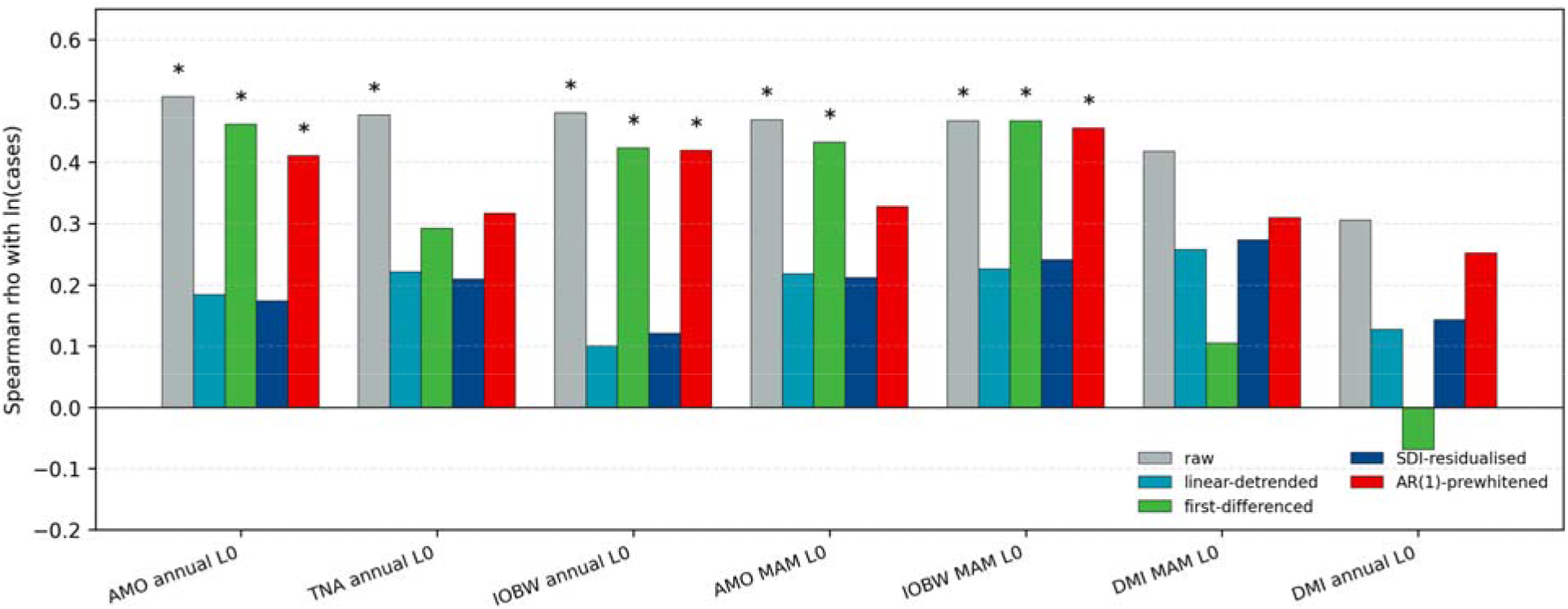
Five-lens trend-correction comparison for the seven zero-lag combinations of the four indices whose raw associations were BH-FDR-significant (AMO, TNA, IOBW and DMI). Bars show Spearman rho with ln(cases) under each lens: raw, linear-detrended, first-differenced, SDI-residualised, and AR(1)-prewhitened. AR(1) prewhitening is shown for every predictor. Asterisks mark BH-FDR significance at 5 percent for the raw, linear-detrended, first-differenced and SDI-residualised lenses, and a bootstrap 95 percent confidence interval excluding zero for the AR(1)-prewhitened lens. The annual AMO and both the annual and spring IOBW index remain significant under first-differencing and AR(1) prewhitening but not under linear detrending or SDI residualisation, locating their associations in inter-annual variation that is still partly entangled with the shared trend.

**Figure 5.**
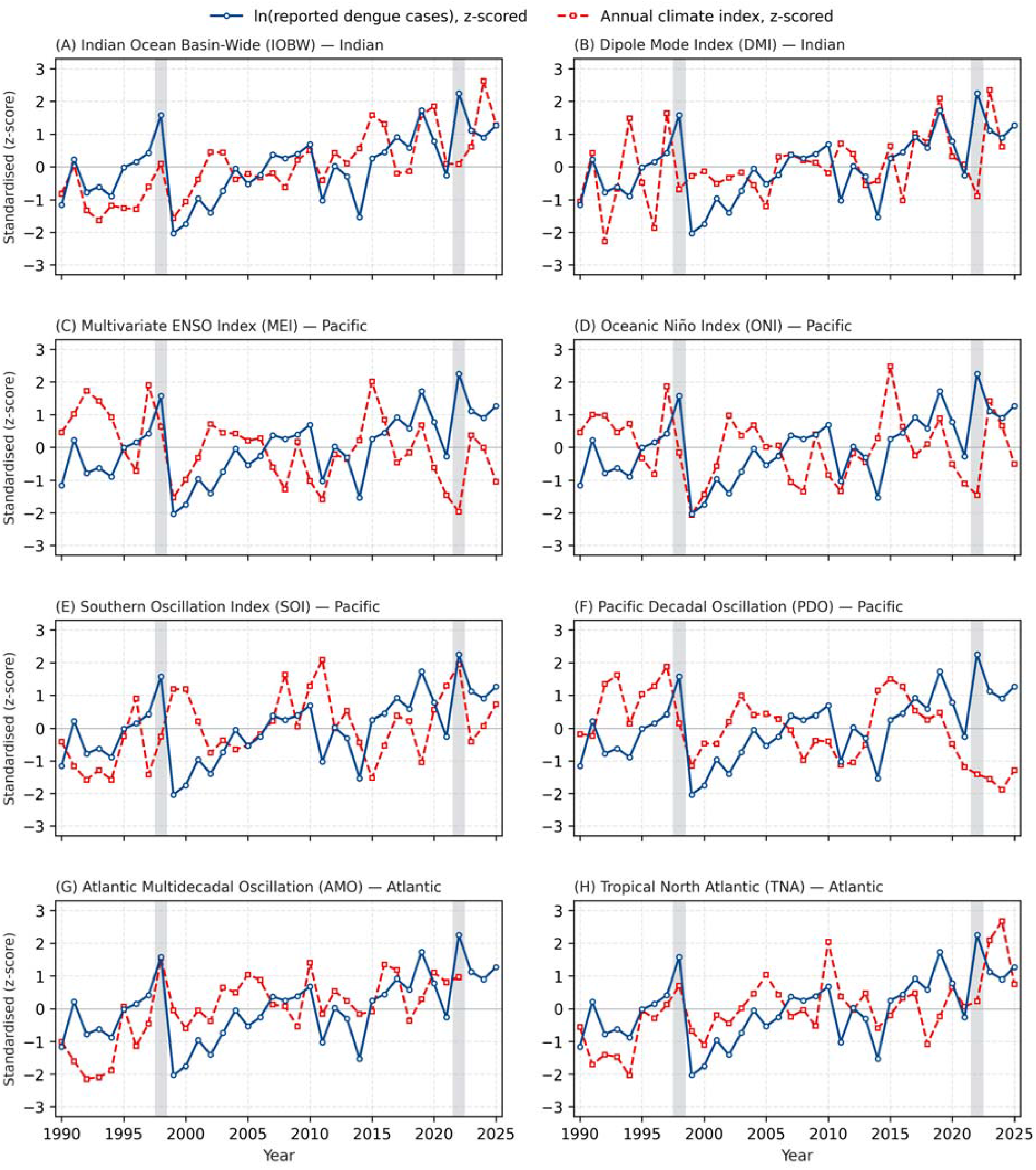
Vietnam’s annual reported dengue cases plotted against all eight tropical sea surface temperature indices, 1990 to 2025, each standardised to a common z-score axis (mean 0, SD 1 over the analysis window). Blue solid line: ln(reported dengue cases). Red dashed line: the annual mean of each index. Panels are grouped by basin: (A, B) Indian Ocean; (C-F) Pacific and El Niño-Southern Oscillation; (G, H) Atlantic. Grey shaded bands mark the 1998 and 2022 outbreak years (detrended log-case z-score > +1·5 SD). Spring (MAM) means track the annual means closely (Supplementary Figure S1) and give the same picture. Visual inspection lets the reader judge co-movement and outbreak alignment directly: the Indian Ocean and Atlantic indices rise and fall with the case record, whereas the Pacific and ENSO indices do not. The corresponding rank correlations, and their behaviour under trend correction, are given in Figure 3 and Figure 4.

For spring IOBW warming, the raw slope is inflated by the 36-year warming trend shared with the case record; after first-differencing it falls but its bootstrap confidence interval still excludes zero - a real inter-annual coupling that also survived AR(1) prewhitening (Supplementary Table S4). In absolute terms, the raw ordinary-least-squares slope linking spring IOBW to log cases was +1·11 per degree Celsius (bootstrap 95 percent confidence interval +0·59 to +1·74); first-differencing reduced it to +0·90 (+0·22 to +1·66), a confidence interval that still excludes zero (Supplementary Figure S3). The bootstrap slope estimate stayed directionally consistent across 13 model specifications, ten of which retained a strictly positive slope confidence interval (Supplementary Figure S4).

## Discussion

Vietnam’s reported dengue burden grew at about 3·5 percent per year over 1990-2025, with 1998 and 2022 classified as outbreak years. Naive rank correlation flagged seven tropical Atlantic and Indian Ocean SST indices as significant predictors, but this finding did not persist following linear detrending. First-differencing preserved signals in both ocean basins; annual AMO and the annual and spring Indian Ocean Basin-Wide index also survived AR(1) prewhitening, whereas spring TNA did not - and ENSO indices failed on every configuration. We attribute this pattern to a multidecadal co-trend between tropical basin warming and Vietnam’s growing surveillance record, together with smaller Atlantic-centred inter-annual signals that the present annual, national-scale data cannot resolve sharply; we do not interpret it as evidence against an underlying climate-dengue link at finer scales. Taken together, most of the apparent climate-dengue association at the national annual scale reflects a shared long-term trend, but a small genuine inter-annual signal – the Indian Ocean Basin-Wide index and the annual Atlantic Multidecadal Oscillation – persists under both trend- and persistence-based correction. Five points follow. First, most candidate climate variables show little evidence of a relationship with dengue at this scale: the Pacific and ENSO indices (MEI, ONI, SOI, PDO) are weak under every configuration, although this may reflect the sub-annual timescale on which ENSO acts rather than a true absence of influence. Second, only a small number of variables – most consistently IOBW and the annual AMO – show associations that persist across lenses. Third, part of the apparent association is attributable to the shared long-term trend, as the leading raw correlations weaken markedly, and several disappear, once that trend is removed by linear detrending or SDI residualisation. Fourth, however, trend is probably not the whole story: were these associations driven entirely by a shared linear trend, they should weaken under both trend-removing and persistence-removing transformations, yet annual and spring IOBW and annual AMO remain distinguishable under both first-differencing and AR(1) prewhitening, indicating a genuine, if modest, inter-annual component alongside the shared trend – or lower-frequency structure that annual data cannot fully separate. Fifth, with only 36 aggregated annual observations the data are too sparse to resolve this further; distinguishing genuine coupling from residual trend, and identifying the timescale on which it acts, will require finer temporal and spatial resolution.

The poor performance of the Pacific and ENSO indices is the first pattern that calls for explanation. Their failure here contrasts with Tian et al., who attributed a large share of regional dengue variance to ENSO using monthly, distributed-lag models^8^. Our annual averaging removes the sub-annual lags through which that signal acts, so the null is a matter of temporal scale rather than evidence against an ENSO-dengue link.

The Indian Ocean and Atlantic indices, by contrast, did register an effect, though a closely qualified one that held under only some lenses. The annual AMO and the Indian Ocean basin-wide index are the clearest cases: both remained statistically distinguishable under first-differencing and persistence-based (AR(1)) prewhitening but not the more aggressive linear detrending or SDI residualisation, indicating a genuine but modest inter-annual coupling that remains partly entangled with the shared multidecadal trend rather than a signal attributable to that trend alone. No direct annual-scale mechanism is obvious, and we do not recommend operationalising it on this evidence. The spring TNA association was not robust under prewhitening and is hypothesis-generating only. Spring IOBW warming retained a first-differenced coupling that also survived AR(1) prewhitening (Supplementary Table S4); a same-year biological pathway - warmer Indian Ocean SSTs lengthening the Aedes breeding window and shortening the extrinsic incubation period - remains plausible^17,18^, but modest in effect.

These contrasts show why the trend-correction approaches are not interchangeable. Linear detrending and SDI residualisation strip the long-term trend, whereas AR(1) prewhitening^19^ removes only short-term serial dependence; a predictor that survives prewhitening but not residualisation has its association co-located with the trend, not with year-to-year variation. This also qualifies how SDI residualisation should be read: because Vietnam’s Socio-demographic Index rises monotonically across 1990 to 2023, residualising on it behaves much like a non-linear detrend, so a signal it removes may reflect the stripping of a monotonic co-trend as much as genuine socioeconomic confounding. Reporting only one trend-correction method risks both false positives (raw correlation alone) and unhelpful nulls (detrending alone), whereas a panel of approaches turns their disagreement into evidence about where a signal sits.

Several important limitations of the analysis should be noted. Annual resolution is intrinsically coarse relative to dengue’s sub-annual transmission dynamics; many biologically important signals at 3-to-6-month lags are averaged out. This constraint is particularly acute for ENSO indices, whose influence on dengue transmission is thought to operate most strongly at sub-annual lags of a few months, so annual averaging may attenuate any genuine ENSO signal. National aggregation obscures sub-national heterogeneity; Vietnam has a substantial north-to-south climate gradient and varying *Aedes* habitat suitability. Source heterogeneity in the merged annual series, particularly for 2011, 2020, 2021-2022, and 2023-2025 (with 2024 derived from a stated 28·2 percent year-over-year change), introduces isolated single-source data points whose comparability with the rest of the series cannot be fully verified. Reported case counts reflect surveillance intensity as well as biological incidence; Vietnam’s surveillance system has grown over the analysis period, and this growth confounds any interpretation of long-term trend.

We did not directly measure the teleconnection pathway; ERA5 reanalysis^20^ or station-level Vietnamese temperature and humidity data would allow a mediation check for the SST to local-climate to dengue chain. A further limitation is Vietnam’s pronounced climate heterogeneity: the northern, central, and southern regions experience substantially different weather patterns, compounded by coastal-inland and microclimatic variation across mountainous and lowland areas. Annual national aggregation masks this spatial heterogeneity, which may partly explain why interannual climate signals are attenuated at the national scale. Because the subtropical north and the year-round tropical south can respond to the same ocean anomaly in opposite directions, and at different times of year, regional signals may partly offset one another in the national total; national aggregation therefore plausibly cancels part of any true coupling rather than merely diluting it.

Set against these limitations, the analysis has several strengths. It draws on an unusually long 36-year national record, which provides the temporal span needed to separate a multidecadal co-trend from genuine year-to-year coupling - a distinction that shorter series cannot support. Rather than relying on a single analytic choice, it applies five complementary trend-correction approaches, so that the behaviour of each candidate association across raw correlation, linear detrending, first-differencing, SDI residualisation and AR(1) prewhitening becomes itself a diagnostic of where the signal sits. Every case record in the merged series carries an explicit labelled source. Bootstrap confidence intervals, multiple-testing correction and a 13-specification robustness check further guard against over-interpreting a single fragile coefficient. Together these features make the conclusions about which climate signals are real, and at what timescale, more robust than a conventional single-transformation correlation analysis would allow.

Four practical implications follow for Vietnam’s operational dengue surveillance and for climate-disease research more broadly. Because this inflation of raw correlations by a shared trend is a property of the data structure rather than of dengue biology, the same panel-of-trend-corrections approach applies to other climate-sensitive mosquito-borne diseases, and extending it across diseases and settings is a natural next step. First, climate-informed analyses drawing on decadal-plus data should report at least two trend-correction approaches alongside the raw correlation, and treat their disagreement as a substantive signal about where the association sits. Second, every dengue case record in national surveillance should retain explicit source provenance (source identifier, case definition, reporting and revision dates) to enable trend-corrected re-analysis. Third, raw rank correlations spanning more than two decades should be interpreted with explicit trend decomposition before any operational use; the AMO-dengue association is a cautionary case in point, and we do not recommend operationalising it for Vietnamese dengue surveillance on the basis of this annual-resolution analysis alone. More generally, we caution against translating raw multi-decadal SST-dengue correlations into operational early-warning products: at this resolution such correlations index a shared secular trend more than any forecastable inter-annual signal, and establishing predictive skill together with a physical mechanism will require sub-national and sub-annual data. An operationally useful early-warning indicator remains a reasonable goal for the field; our results simply locate it at monthly, provincial resolution rather than in national annual correlations. Fourth, the climate-dengue question for Vietnam should be reframed at monthly resolution and at provincial scale, where statistical power and physical mechanism are jointly available.

## Conclusion

At the national annual scale, most large-scale climate indices show little evidence of a relationship with Vietnam’s dengue incidence, and the associations that do appear are concentrated in a few Indian Ocean and Atlantic indices – most consistently the Indian Ocean Basin-Wide index and the annual Atlantic Multidecadal Oscillation. Vietnam’s reported dengue burden rose at about 3·5 percent per year over 1990-2025, a trend that reflects population growth and surveillance strengthening as much as any underlying biological change. Under the primary analysis (first-differencing), signals in both ocean basins persisted; the annual AMO and the annual and spring Indian Ocean basin-wide index also survived AR(1) prewhitening, while the spring TNA signal did not; the remaining four approaches, treated as sensitivity analyses, broadly agree on this pattern. We were therefore unable to demonstrate that climate is the primary driver of national annual dengue: much of the raw association is consistent with a shared long-term trend rather than strong year-to-year coupling, though a small genuine inter-annual signal (annual AMO and the Indian Ocean basin-wide index) persists under both trend- and persistence-based correction. We recommend that climate-disease analyses spanning more than two decades report a panel of trend-correction approaches, treat their disagreement as diagnostic, and reserve any forecasting claims for predictors that persist under both trend- and persistence-based checks - and even then, only at finer temporal and spatial resolution than the national annual scale analysed here.

The policy relevance of these distinctions is sharpened by the current climate context. The World Meteorological Organization’s State of the Climate in Asia 2025 report documents record ocean-heat content across the Indian Ocean and Western Pacific and a basin-wide transition from La Niña toward El Niño during 2026 ^21^. Because both regional sea surface temperature and Vietnam’s reported dengue burden are rising on shared multidecadal trajectories, analyses keyed to raw long-span SST-dengue correlations risk attributing this common trend to a genuine climate signal - the very misattribution our five-lens framework is designed to expose. Three operational implications follow: climate-informed dengue forecasts should be validated on inter-annual skill rather than hindcast fit to a co-trend; the onset of El Niño, which carries no robust inter-annual dengue signal in our Vietnam analysis, should not by itself trigger resource mobilisation; and vector-control and surveillance capacity - strengthened but unevenly sustained since COVID-19 - should remain the primary line of outbreak preparedness rather than be deferred to climate prediction.

## Authors’ contributions

Doanh Nguyen-Ngoc: Conceptualization, Methodology, Funding acquisition, Supervision, Project administration, Writing – original draft, Writing – review & editing. James M. Trauer: Methodology, Validation, Writing – review & editing. Hai Son Vo: Resources, Writing – review & editing. Andrew W. Taylor-Robinson: Methodology, Writing – review & editing. Thanh H. Nguyen: Writing – review & editing. Viet Long Bui: Methodology, Data curation, Software, Formal analysis, Visualization, Writing – original draft, Writing – review & editing. All authors read and approved the final version of the manuscript.

## Data sharing

All data analysed in this study are publicly available. Vietnam’s annual dengue case counts were obtained from OpenDengue version 1.3 (https://opendengue.org) and from publicly released Vietnam Ministry of Health figures. Global Burden of Disease 2023 dengue estimates and population denominators were retrieved from the Institute for Health Metrics and Evaluation Global Burden of Disease Results Tool (https://vizhub.healthdata.org/gbd-results/). Tropical sea surface temperature indices were obtained from the National Oceanic and Atmospheric Administration Physical Sciences Laboratory (https://psl.noaa.gov/data/climateindices/) and the National Centers for Environmental Information. The analysis code that reproduces all results and figures are available on: https://github.com/vlbui/dengue-climate-vietnam.

## Ethics

This study used only aggregated, de-identified, publicly available secondary data and did not involve human participants, identifiable personal information, or animal subjects. Ethics committee approval and informed consent were therefore not required.

## Acknowledgements

We thank the Project Tycho, WHO Western Pacific Regional Office, and Vietnam Ministry of Health teams whose surveillance data contributions are consolidated in OpenDengue V1·3 and whose 2023 to 2025 public communications underpin this analysis. We thank the NOAA Physical Sciences Laboratory and NOAA National Centers for Environmental Information for making tropical SST indices publicly available.

