## Supplementary Material for "Is climate a primary driver of Vietnam’s dengue, or a shared long-term trend? A five-method analysis over 36 years"

**Supplementary Materials**

This document contains Supplementary Methods S1 and S2, Supplementary Figures S1 to S5, and Supplementary Tables S1 to S4. Section numbers are referenced from the main text; literature references are listed separately at the end of this document and are numbered independently of the main manuscript.

### S1. Supplementary Methods

#### S1.1 Dengue case series and source merge rule

The national annual dengue series for 1990-2025 was assembled from the OpenDengue database (version 1.3)^1^ for 1990-2022 and from Vietnamese Ministry of Health communications for 2023-2025. The OpenDengue release provides three extracts for Vietnam (national, temporal, and spatio-temporal); cross-validation over the 1994-2010 overlap window confirmed an exact 1·000 ratio between summed monthly counts and native annual counts.

Within each year a single source was selected by a fixed-priority rule. For most years the first available of the following was used, in order: (1) World Health Organization Western Pacific Regional Office records^2^, used when the year had at least 48 weekly or at least 11 monthly rows, with sub-annual counts summed to an annual total; (2) the Project Tycho^3^ national annual record; (3) the multi-country literature compilation. Four years used a designated Vietnam-specific source: 2011 used Vietnam monthly literature records summed to an annual total, 2020 used the Vietnamese Ministry of Health annual report, and 2021 and 2022 used Vietnam literature annual records. Each year was therefore assigned exactly one labelled source; the source assigned to every year is listed in Supplementary Table S1.

#### S1.2 Data extraction protocol

This section records the retrieval procedure in full so the dataset can be reconstructed. Abbreviations are spelled out where they first appear.

**Step 1 - dengue cases, 1990-2022.** The OpenDengue database version 1.3 was downloaded from opendengue.org on 19 April 2026. Three extracts (national, temporal, and spatio-temporal) were filtered to Vietnam using the country code VNM.

**Step 2 - dengue cases, 2023-2025.** Values were taken from public Ministry of Health (MoH) communications: 172,000 cases for 2023, 148,237 for 2024 (derived as 190,040 divided by 1·282, the stated year-on-year change), and 190,040 cases for 2025.

**Step 3 - Socio-demographic Index.** The Vietnamese Socio-demographic Index (SDI) series was taken from the IHME covariates release^4^.

**Step 4 - climate.** Eight tropical sea surface temperature (SST) indices were selected as candidate predictors, spanning three ocean basins whose SST modes are implicated in Southeast Asian monsoon variability and regional climate. The dengue-specific evidence motivating each basin is given in the Introduction of the main text; this section provides index definitions, data sources, and processing.

**Indian Ocean.** The Indian Ocean Basin-Wide index (IOBW) and the Dipole Mode Index (DMI) capture the basin-averaged SST and the zonal SST gradient (Indian Ocean Dipole) respectively. Chen et al. identified annual IOBW as the single strongest predictor of dengue across 46 countries, outperforming ENSO diagnostics^5^. DMI, retrieved from the National Oceanic and Atmospheric Administration (NOAA) Physical Sciences Laboratory, has been linked to monsoon-onset anomalies and Southeast Asian precipitation shifts in positive-IOD years ^6^. IOBW was computed locally from the ERSST v5 gridded product as anomalies relative to a 1971-2000 climatological baseline^7^

.

**Pacific and ENSO.** The Multivariate ENSO Index version 2 (MEI) ^8^, the Oceanic Niño Index (ONI), and the Southern Oscillation Index (SOI)^9^ were included as complementary ENSO diagnostics. Cazelles et al.^10^ reported a non-stationary ENSO influence on synchronised dengue epidemics in Thailand at monthly resolution, and Chen and colleagues ^5^ attributed 63 percent of dengue variance in 57 countries to ENSO in a monthly distributed-lag non-linear model with 3 to 6 month lags. The Pacific Decadal Oscillation (PDO)^11^ modulates the background state on which ENSO teleconnections propagate and has been discussed in Asia-Pacific vector-borne disease contexts^12^. MEI, SOI, ONI and PDO were obtained from NOAA PSL, NOAA CPC, and NOAA NCEI.

**Atlantic.** The Atlantic Multidecadal Oscillation (AMO)^13^ and the Tropical North Atlantic index (TNA) are candidate trans-basin teleconnection sources that can modulate the Pacific Walker cell and, in turn, Southeast Asian monsoon delivery timing. AMO has been linked to continental-US precipitation patterns and a limited number of vector-borne disease studies; dengue-specific literature on AMO and TNA remains sparse, so we included them as exploratory predictors. AMO and TNA were retrieved from NOAA PSL.

Each index was aggregated to an annual mean and to a March-to-May (MAM) spring mean, the latter following Chen and colleagues, who reported spring IOBW as the globally dominant annual dengue predictor. This yields 16 candidate predictors aligned to 36 annual dengue observations. All indices provide full 1990 to 2025 monthly coverage except AMO and DMI. A year-level quality filter retains only years with at least 11 monthly observations, so AMO ends in 2022 (2023 contributes a single month) and DMI ends in 2024 (2025 contributes four months); both therefore have slightly reduced samples, and all tables and figures use the filtered series.

#### S1.3 Statistical methods

Trend in the annual case series was tested with the Mann-Kendall test^14,15^ using tie-corrected variance and a continuity correction, with the Sen slope^16^ as the accompanying estimate. Associations between dengue and each climate index were measured with the Spearman rank correlation coefficient^17^, with two-sided p-values from a Student t approximation. To control the false discovery rate across multiple predictors, p-values were adjusted by the Benjamini-Hochberg procedure \^18^ within each lag. Where an effect size was reported, a 95 percent confidence interval was obtained from 5,000 paired bootstrap resamples.

#### S1.4 Trend-correction procedures

Two trend-correction transforms were applied to the log-transformed case series and to each climate index before the Spearman correlation was recomputed. Linear detrending removes the fitted linear trend: for a series s observed at times t, an ordinary least squares (OLS) line is fitted and subtracted,

$$r_{t} = s_{t} - (\beta t + \alpha)$$

where β and α are the fitted slope and intercept. First-differencing removes the trend by taking year-on-year changes,

$$\Delta s_{t} = s_{t} - s_{t-1}$$

for t = 2 to N.

#### S1.5 AR(1) prewhitening procedure

The log-transformed case series for 1990-2023 was modelled as a first-order autoregressive, AR(1) ^19^, process,

$$ln({cases}_{t}) = c + \varphi ln({cases}_{t-1}) + \varepsilon_{t}$$

where c is a constant, φ is the autoregressive coefficient, and the final term is the residual. The fitted φ was approximately +0·34. The prewhitened residual series was then correlated, by Spearman rank, with each climate predictor at lags of zero and one year. For all six predictors that survived first-differencing (annual and spring AMO and IOBW at lag zero, and annual and spring TNA at a one-year lag), percentile bootstrap 95 percent confidence intervals on the prewhitened correlation were computed from 1,000 paired resamples.

AR(1) prewhitening removes first-order serial dependence from the dengue series but does not remove the multidecadal trend that the AMO shares with the growth of Vietnamese surveillance. This is what makes the procedure informative when its result is contrasted with Socio-demographic Index residualisation, which removes that trend explicitly: a predictor that survives prewhitening but not residualisation has its association located in the shared trend rather than in year-to-year coupling. Because the Vietnamese Socio-demographic Index rises monotonically over 1990 to 2023, residualising on a quadratic SDI fit behaves similarly to non-linear (quadratic) detrending; a signal removed by SDI residualisation may therefore reflect the stripping of a monotonic co-trend as much as genuine socioeconomic confounding, and we interpret it accordingly.

#### S1.6 Software

All analyses were run in Python ^20^. The full dependency list is in the project repository environment file.

### S2. Additional results

#### S2.1 Supplementary Figures

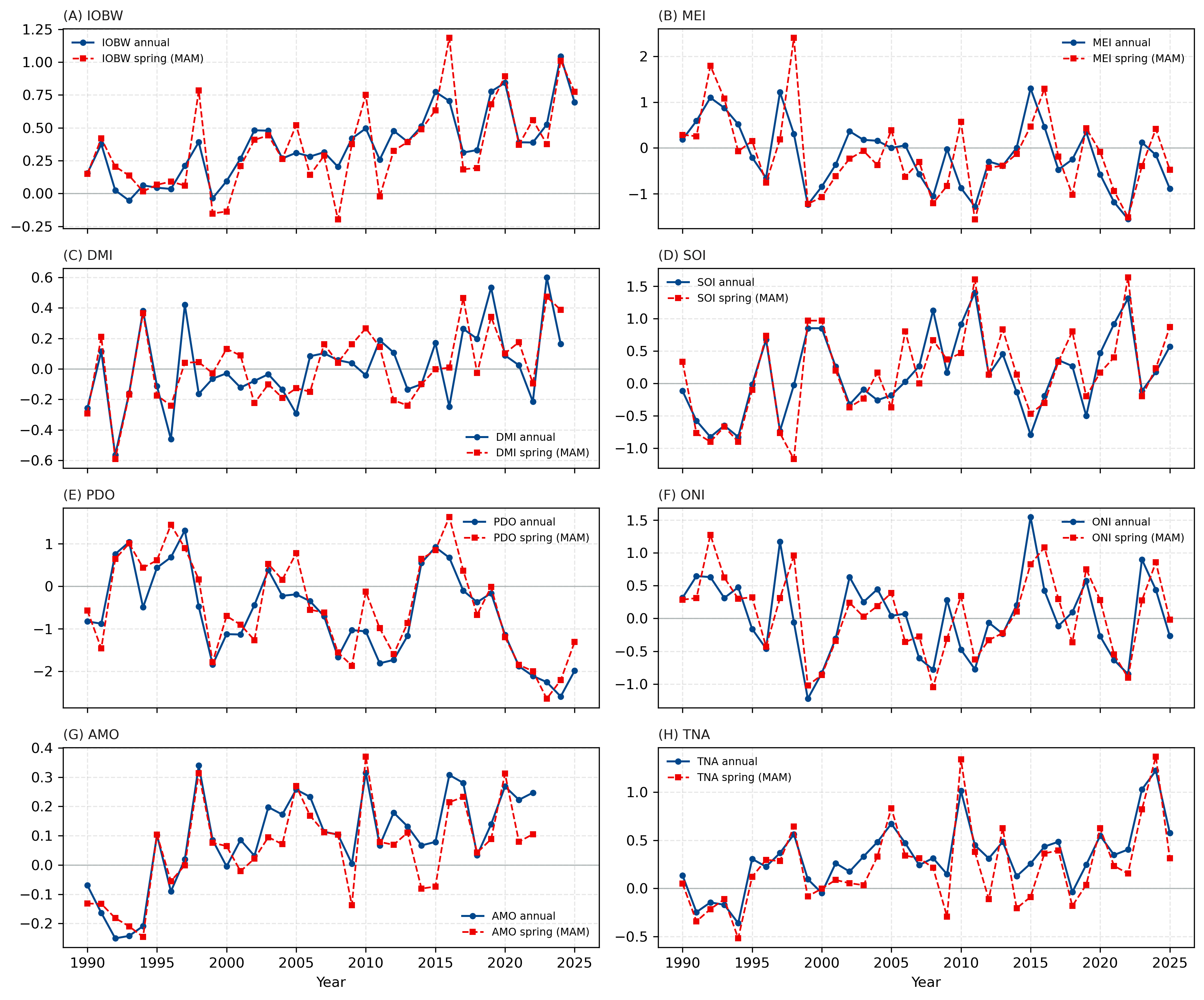

**Figure S1.** Annual and spring (MAM) means of eight tropical SST indices over 1990 to 2025. Solid blue line with circles: annual mean. Dashed red line with squares: March to May spring mean. Zero reference line is drawn on each panel.

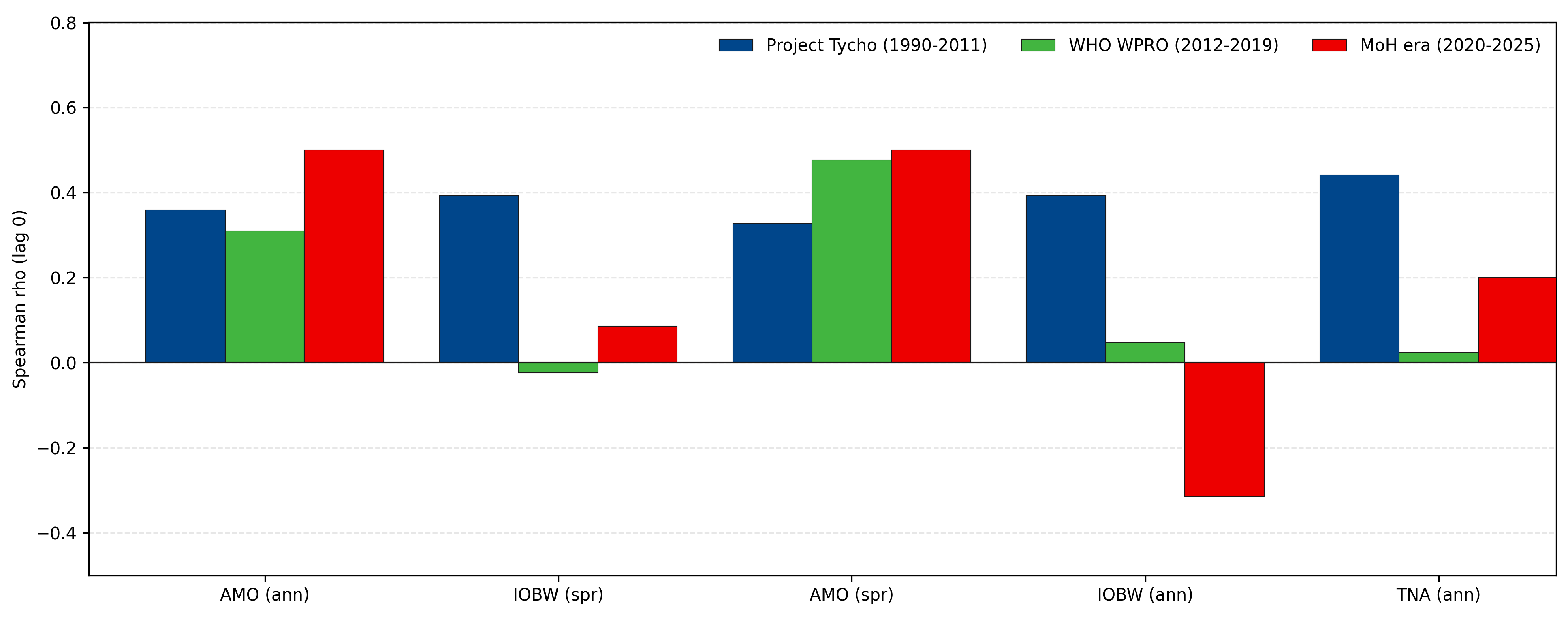

**Figure S2.** Period-stratified robustness. Bars show lag-0 Spearman rho within three periods defined by data source - Project Tycho (1990-2011), WHO WPRO (2012-2019) and Ministry of Health (2020-2025) - for the five top primary predictors. Splitting by data origin tests whether the associations shift at a source handover: the leading IOBW signal still weakens after the earliest period, while AMO stays positive across sources. The Ministry of Health period is short (n = 6; n = 3 for AMO, which ends in 2022), so those bars are interpreted with caution.

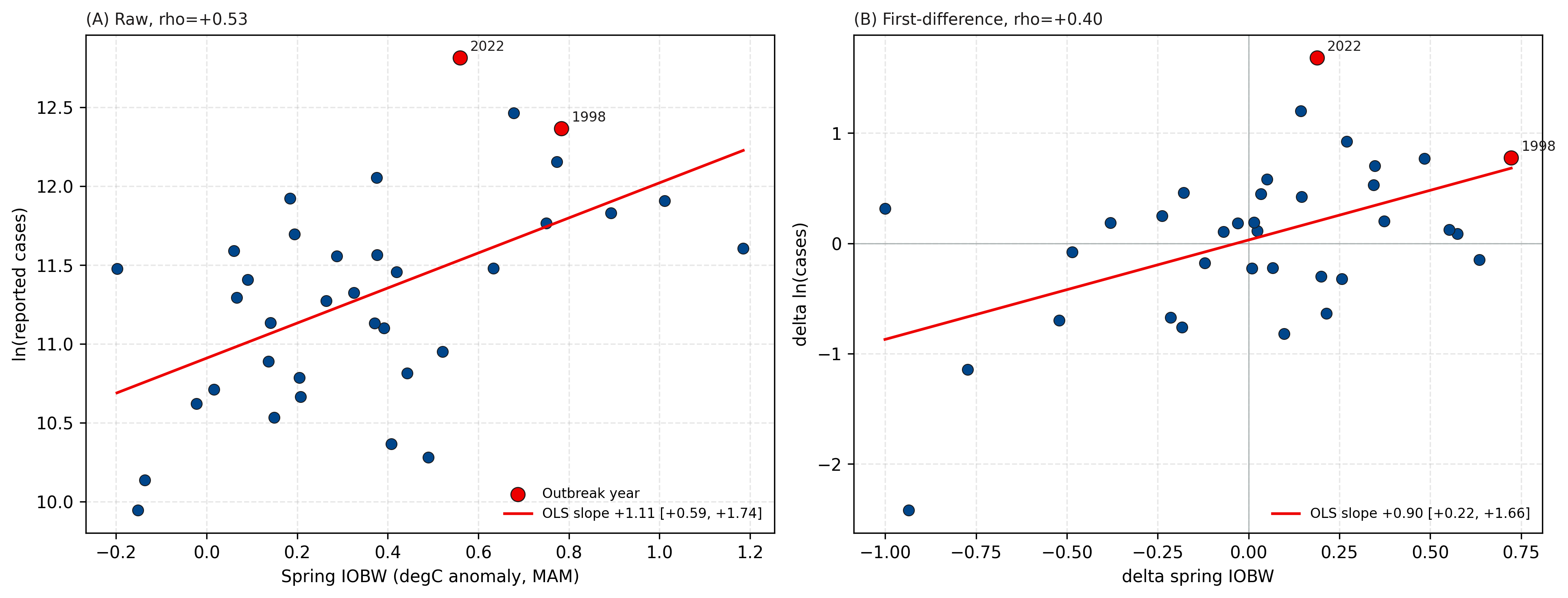

**Figure S3.** IOBW-focused two-panel scatter. (A) Raw spring IOBW (March-May mean) versus ln(reported cases). (B) First-difference scatter. Red markers and year labels indicate the two outbreak years defined in main-text Methods and Figure 2 (detrended log-case z-score above +1·5 standard deviations: 1998 and 2022).

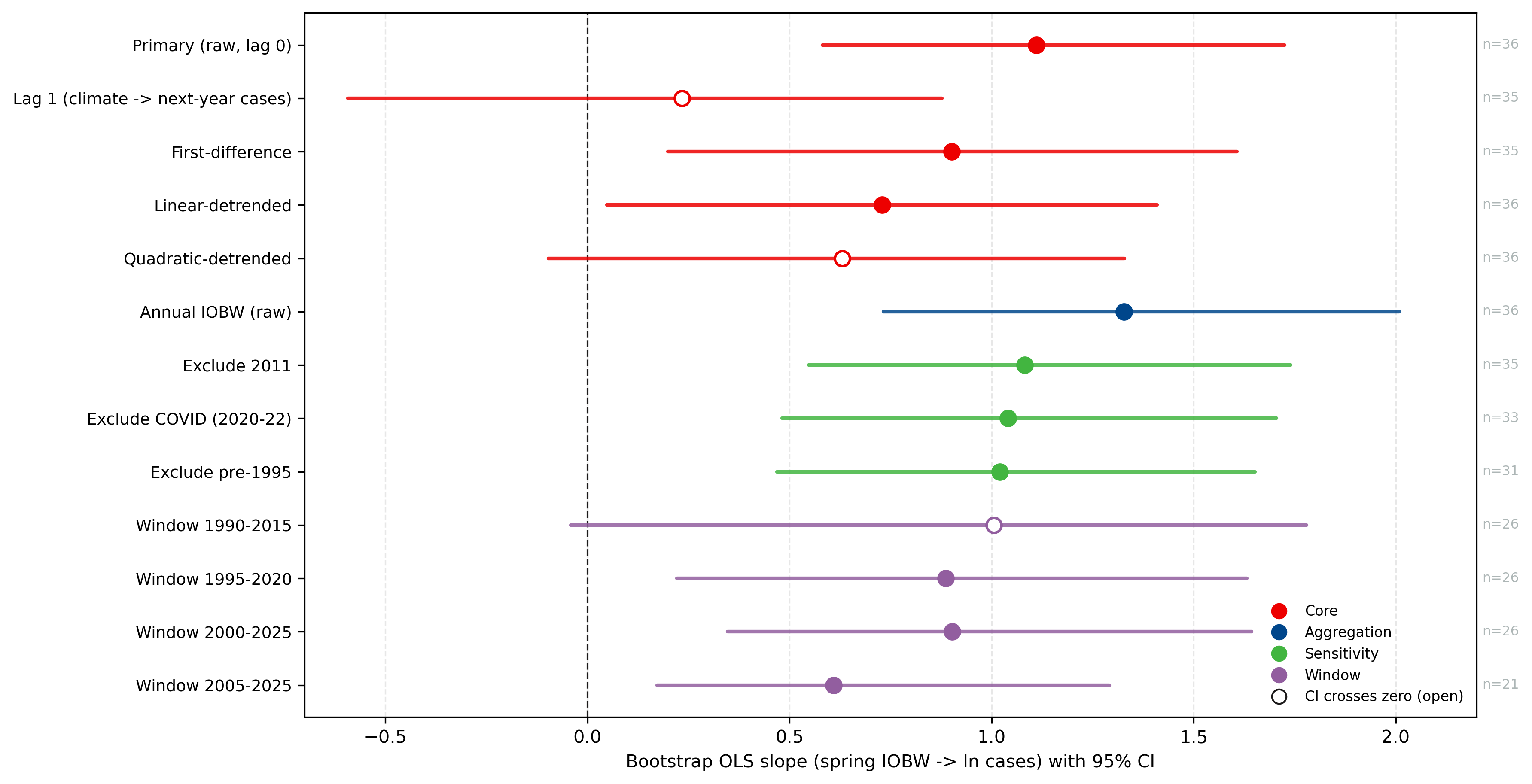

**Figure S4.** Bootstrap OLS slope (spring IOBW to ln-cases) with 95 percent CI across 13 specifications, grouped as Core, Aggregation, Sensitivity, and Window. Filled circles: CI excludes zero. Open circles: CI crosses zero. Ten of thirteen specifications show a strictly positive slope CI.

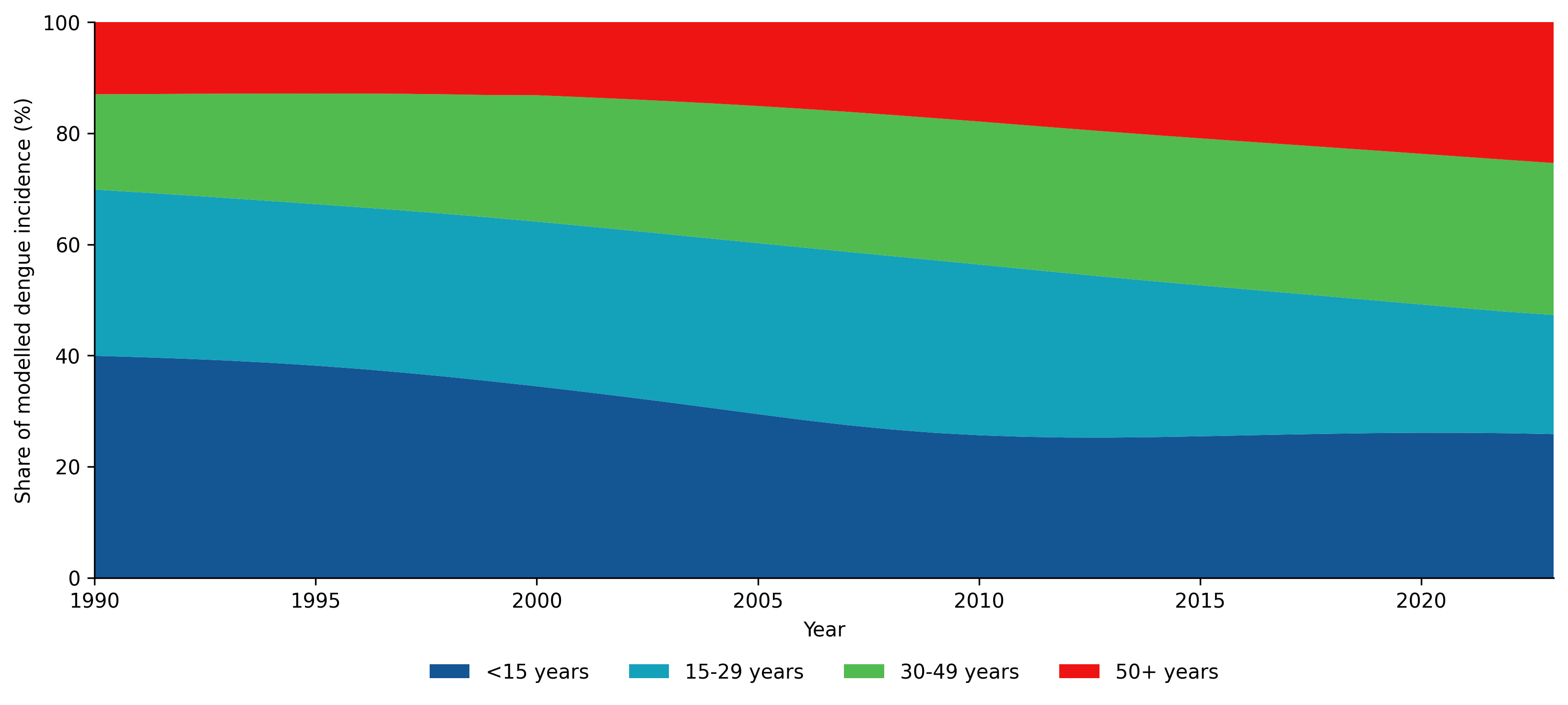

**Figure S5.** Age distribution of dengue burden in Vietnam, 1990 to 2023. Share of Global Burden of Disease 2023 modelled dengue incidence by broad age band, presented as descriptive context only and not used in the primary climate analysis. The share among children younger than 15 years declined from about 40 percent to 26 percent, while the share among adults aged 50 years and older rose from about 13 percent to 25 percent over the period.

#### S2.2 Supplementary Tables

**Table S1. Source assigned to each year of the dengue series**

| **Year** | **Reported cases** | **Assigned source** |
| --- | --- | --- |
| 1990 | 37,569 | Project Tycho |
| 1991 | 94,630 | Project Tycho |
| 1992 | 48,333 | Project Tycho |
| 1993 | 53,674 | Project Tycho |
| 1994 | 44,944 | Project Tycho |
| 1995 | 80,447 | Project Tycho |
| 1996 | 89,963 | Project Tycho |
| 1997 | 108,000 | Project Tycho |
| 1998 | 234,866 | Project Tycho |
| 1999 | 20,861 | Project Tycho |
| 2000 | 25,269 | Project Tycho |
| 2001 | 42,878 | Project Tycho |
| 2002 | 31,760 | Project Tycho |
| 2003 | 49,751 | Project Tycho |
| 2004 | 78,692 | Project Tycho |
| 2005 | 56,980 | Project Tycho |
| 2006 | 68,532 | Project Tycho |
| 2007 | 104,393 | Project Tycho |
| 2008 | 96,451 | Project Tycho |
| 2009 | 105,370 | Project Tycho |
| 2010 | 128,831 | Project Tycho |
| 2011 | 40,998 | Literature (Vietnam) |
| 2012 | 82,838 | WHO WPRO |
| 2013 | 66,320 | WHO WPRO |
| 2014 | 29,181 | WHO WPRO |
| 2015 | 96,751 | WHO WPRO |
| 2016 | 109,649 | WHO WPRO |
| 2017 | 150,502 | WHO WPRO |
| 2018 | 120,163 | WHO WPRO |
| 2019 | 259,070 | WHO WPRO |
| 2020 | 137,470 | MoH annual report |
| 2021 | 68,268 | Literature (Vietnam) |
| 2022 | 367,729 | Literature (Vietnam) |
| 2023 | 172,000 | MoH communication |
| 2024 | 148,237 | MoH communication |
| 2025 | 190,040 | MoH communication |

**Table S2. Raw Spearman correlations for the 16 climate predictors**

| **Index** | **Season** | **Lag (yr)** | **n** | **Spearman rho** | **p-value** | **BH-adjusted p** | **Significant** |
| --- | --- | --- | --- | --- | --- | --- | --- |
| IOBW | Annual | 0 | 36 | +0·534 | <0·001 | 0·001 | Yes |
| TNA | Annual | 0 | 36 | +0·530 | <0·001 | 0·001 | Yes |
| IOBW | Spring (MAM) | 0 | 36 | +0·529 | <0·001 | 0·001 | Yes |
| AMO | Annual | 0 | 33 | +0·506 | 0·001 | 0·004 | Yes |
| AMO | Spring (MAM) | 0 | 33 | +0·469 | 0·003 | 0·009 | Yes |
| DMI | Spring (MAM) | 0 | 35 | +0·456 | 0·003 | 0·009 | Yes |
| DMI | Annual | 1 | 36 | +0·425 | 0·006 | 0·014 | Yes |
| TNA | Spring (MAM) | 0 | 36 | +0·411 | 0·009 | 0·017 | Yes |
| ONI | Spring (MAM) | 0 | 36 | +0·264 | 0·111 | 0·197 | No |
| ONI | Annual | 1 | 36 | +0·222 | 0·184 | 0·294 | No |
| MEI | Spring (MAM) | 0 | 36 | +0·172 | 0·309 | 0·449 | No |
| PDO | Spring (MAM) | 0 | 36 | -0·149 | 0·380 | 0·507 | No |
| PDO | Annual | 0 | 36 | -0·132 | 0·437 | 0·538 | No |
| SOI | Annual | 0 | 36 | +0·109 | 0·521 | 0·595 | No |
| MEI | Annual | 0 | 36 | -0·054 | 0·755 | 0·766 | No |
| SOI | Spring (MAM) | 1 | 36 | +0·051 | 0·766 | 0·766 | No |

**Table S3. Trend-correction comparison across configurations***Note. Raw Spearman coefficients in this table are computed on the common analysis window shared by all four trend-correction methods (constrained by Socio-demographic Index covariate coverage, 1990–2023; n = 34, or n = 33 for AMO), so the four lenses are compared on identical samples; they therefore differ slightly from the full-window raw coefficients in Table S2 (n up to 36).*

| **Index** | **Season** | **Lag (yr)** | **Raw** | **Linear-detrend** | **First-difference** | **SDI-residual** |
| --- | --- | --- | --- | --- | --- | --- |
| AMO | Spring (MAM) | 0 | +0·469 | +0·218 | +0·433 | +0·212 |
| AMO | Spring (MAM) | 1 | +0·043 | -0·213 | -0·393 | -0·226 |
| AMO | Annual | 0 | +0·506 | +0·184 | +0·462 | +0·173 |
| AMO | Annual | 1 | +0·105 | -0·213 | -0·308 | -0·220 |
| DMI | Spring (MAM) | 0 | +0·418 | +0·257 | +0·105 | +0·272 |
| DMI | Spring (MAM) | 1 | +0·236 | +0·060 | +0·108 | +0·081 |
| DMI | Annual | 0 | +0·305 | +0·127 | -0·069 | +0·143 |
| DMI | Annual | 1 | +0·364 | +0·185 | +0·374 | +0·206 |
| IOBW | Spring (MAM) | 0 | +0·468 | +0·226 | +0·468 | +0·241 |
| IOBW | Spring (MAM) | 1 | +0·074 | -0·199 | -0·360 | -0·210 |
| IOBW | Annual | 0 | +0·480 | +0·100 | +0·423 | +0·120 |
| IOBW | Annual | 1 | +0·239 | -0·139 | -0·046 | -0·143 |
| MEI | Spring (MAM) | 0 | +0·170 | +0·291 | +0·290 | +0·297 |
| MEI | Spring (MAM) | 1 | -0·110 | -0·028 | -0·186 | -0·035 |
| MEI | Annual | 0 | -0·003 | +0·138 | +0·089 | +0·138 |
| MEI | Annual | 1 | +0·036 | +0·180 | +0·272 | +0·170 |
| ONI | Spring (MAM) | 0 | +0·254 | +0·341 | +0·229 | +0·353 |
| ONI | Spring (MAM) | 1 | -0·048 | -0·008 | -0·097 | -0·006 |
| ONI | Annual | 0 | +0·097 | +0·158 | +0·071 | +0·163 |
| ONI | Annual | 1 | +0·155 | +0·239 | +0·301 | +0·233 |
| PDO | Spring (MAM) | 0 | -0·055 | +0·106 | +0·130 | +0·097 |
| PDO | Spring (MAM) | 1 | +0·092 | +0·195 | +0·004 | +0·186 |
| PDO | Annual | 0 | -0·006 | +0·180 | +0·079 | +0·166 |
| PDO | Annual | 1 | +0·219 | +0·308 | +0·169 | +0·299 |
| SOI | Spring (MAM) | 0 | -0·085 | -0·233 | -0·192 | -0·239 |
| SOI | Spring (MAM) | 1 | +0·056 | -0·070 | -0·057 | -0·055 |
| SOI | Annual | 0 | +0·075 | -0·084 | -0·114 | -0·081 |
| SOI | Annual | 1 | -0·009 | -0·157 | -0·274 | -0·146 |
| TNA | Spring (MAM) | 0 | +0·370 | +0·207 | +0·204 | +0·196 |
| TNA | Spring (MAM) | 1 | +0·005 | -0·189 | -0·452 | -0·196 |
| TNA | Annual | 0 | +0·477 | +0·221 | +0·291 | +0·209 |
| TNA | Annual | 1 | +0·076 | -0·204 | -0·438 | -0·210 |

**Table S4. AR(1)-prewhitened correlations for the surviving predictors**

Spearman correlation between each climate predictor and the AR(1)-prewhitened dengue residual series, for the six predictors that survived first-differencing and, for completeness, for the remaining zero-lag predictors shown in Figure 4. The 95 percent confidence interval (CI) is a percentile bootstrap from 1,000 paired resamples.

| **Index** | **Season** | **Lag (yr)** | **n** | **Prewhitened rho** | **95% CI lower** | **95% CI upper** |
| --- | --- | --- | --- | --- | --- | --- |
| AMO | Annual | 0 | 32 | +0·411 | +0·079 | +0·653 |
| TNA | Spring (MAM) | 1 | 33 | -0·165 | -0·506 | +0·234 |
| IOBW | Spring (MAM) | 0 | 33 | +0·456 | +0·090 | +0·709 |
| IOBW | Annual | 0 | 33 | +0·419 | +0·079 | +0·676 |
| AMO | Spring (MAM) | 0 | 32 | +0·328 | -0·013 | +0·614 |
| TNA | Annual | 1 | 33 | -0·112 | -0·468 | +0·287 |
| TNA | Annual | 0 | 33 | +0·317 | -0·051 | +0·602 |
| DMI | Spring (MAM) | 0 | 33 | +0·309 | -0·053 | +0·573 |
| DMI | Annual | 0 | 33 | +0·252 | -0·115 | +0·572 |
